# Deep Brain Stimulation Normalizes Amygdala Responsivity in Treatment-Resistant Depression

**DOI:** 10.1101/2022.09.08.22279718

**Authors:** N. Runia, I.O. Bergfeld, B.P. de Kwaasteniet, J. Luigjes, J. van Laarhoven, P. Notten, G. Beute, P. van den Munckhof, P.R. Schuurman, D.A.J.P. Denys, G.A. van Wingen

## Abstract

Deep brain stimulation (DBS) of the ventral anterior limb of the internal capsule (vALIC) is a promising intervention for treatment-resistant depression (TRD). However, the working mechanisms of vALIC DBS in TRD remain largely unexplored. As major depressive disorder has been associated with aberrant amygdala functioning, we investigated whether vALIC DBS affects amygdala responsivity and functional connectivity. To investigate the long-term effects of DBS, eleven patients with TRD performed an implicit emotional face viewing paradigm during functional magnetic resonance imaging (fMRI) before DBS surgery and after DBS parameter optimization. Sixteen matched healthy controls performed the fMRI paradigm at two time points to control for test-retest effects. To investigate the short-term effects of DBS de-activation after parameter optimization, thirteen patients additionally performed the fMRI paradigm after double-blind periods of active and sham stimulation. Results showed that TRD patients had decreased right amygdala responsivity compared to healthy controls at baseline. Long-term vALIC DBS normalized right amygdala responsivity, which was associated with faster reaction times. Furthermore, active compared to sham DBS increased amygdala connectivity with sensorimotor and cingulate cortices. These results suggest that vALIC DBS may relieve emotional blunting and depression by restoring amygdala responsivity and behavioral vigilance.

## 1. Introduction

Deep brain stimulation (DBS) is a promising intervention for treatment-resistant depression (TRD). DBS of the ventral anterior limb of the internal capsule (vALIC), a white matter bundle that connects the frontal cortex with subcortical brain regions (1), results in approximately 40% of patients being classified as responders after one year of treatment (2). Additionally, active vALIC DBS significantly reduced depressive symptoms compared to sham stimulation in a double-blind cross-over study (3).

At present, no studies have attempted to determine the working mechanisms of vALIC DBS for TRD using functional neuroimaging techniques. However, results from several positron emission tomography studies targeting the nucleus accumbens (NAc), which is very close to the vALIC target (2), suggest that focal stimulation can have widespread effects on brain metabolism. More specifically, studies show that NAc DBS induced decreased glucose metabolism in the posterior cingulate, caudate nucleus, thalamus, cerebellum and orbital and dorsomedial prefrontal cortex and increased metabolism the NAc and dorsolateral prefrontal cortex (4-6). However, these studies were performed with small sample sizes and without comparison to a control group.

Affective symptoms in depression, such as sustained negative affect and difficulties with experiencing positive affect, are thought to be the result of aberrant emotion reactivity and regulation (7). Neurobiological models of depression highlight the role of a frontolimbic network in emotion dysregulation (8, 9). A meta-analysis of neuroimaging studies showed that several brain regions, including the amygdala, respond more vigorously to negative stimuli in patients with depression compared to healthy controls (10). During processing of facial expressions, decreased functional connectivity between the amygdala and the supragenual/dorsal anterior cingulate cortex (ACC) and dorsolateral prefrontal cortex, and increased functional connectivity between the amygdala and the subgenual ACC was found (11). We therefore hypothesized that vALIC DBS may normalize aberrancies in amygdala responsivity and functional connectivity.

To test this hypothesis, TRD patients receiving vALIC DBS treatment performed an implicit emotional face viewing task that is known to activate the amygdala (12) during functional magnetic resonance imaging (fMRI). Patients underwent fMRI scans before vALIC DBS implantation and after DBS parameter optimization. Matched healthy controls were included and performed the fMRI task at two time points to control for test-retest effects. We then analyzed amygdala responsivity as well as amygdala connectivity with other brain regions to assess network functioning. To assess short-term effects of vALIC DBS de-activation after parameter optimization on amygdala responsivity and connectivity, TRD patients also performed the fMRI task after double-blind periods of active and sham stimulation.

## 2. Methods

### 2.1 Participants

We included patients with TRD as well as healthy controls in a longitudinal study followed by a randomized crossover phase (for patients only). This imaging study was an add-on to a previously reported clinical trial for DBS in TRD (Bergfeld, 2016). The study was approved by the medical ethics boards of the two participating hospitals: Academic Medical Center, Amsterdam [AMC] and St Elisabeth Hospital, Tilburg [SEH]. All included participants provided written informed consent.

At inclusion, patients had to be aged 18-65 years and have a primary diagnosis of major depressive disorder (MDD) according to the DSM-IV (assessed with a semi-structured clinical interview for DSM-IV disorders), an illness duration of more than 2 years, a 17-item Hamilton Depression Rating Scale (HAM-D-17) score of 18 or higher, and a Global Assessment of Function sore of 45 or lower. Additionally, patients had to be treatment-resistant, defined as a failure of at least two different classes of second-generation antidepressants, one trial of a tricyclic antidepressant, one trial of a tricyclic antidepressant with lithium augmentation, one trial of a monoamine oxidase inhibitor, and six or more sessions of bilateral electroconvulsive therapy. Patients who fulfilled the above criteria and remained stable with maintenance electroconvulsive therapy, but relapsed after discontinuation of that therapy, were also eligible. Patients had to be able to understand the consequences of the procedure (IQ >80), and capable of making choices without coercion. Exclusion criteria were: Parkinson’s disease, dementia, epilepsy, bipolar disorder, schizophrenia or history of psychosis unrelated to MDD, antisocial personality disorder, current tic disorder, an organic cause of depression, substance abuse during the past 6 months, unstable physical condition, pregnancy, or general contraindications for surgery.

Healthy controls were matched by age, sex, and education level. They and their first-degree relatives needed to have negative lifetime histories of psychiatric illness.

### 2.2 Treatment

Four-contact leads were implanted bilaterally and connected to a neurostimulator (lead: 3389; stimulator: Activa PC, Medtronic, Minneapolis, MN, USA). The electrodes were implanted with the most ventral contact at the core of the NAc and the three more dorsal contacts in the ventral ALIC. Following a three-week recovery period DBS setting optimization was started. Optimization ended after a stable response was reached for at least four weeks or after a maximum of 52 weeks. Details on the surgery and DBS treatment have been described earlier (3).

### 2.3 Study Design

MRI scanning was performed three weeks prior to DBS surgery (baseline), and after DBS parameter optimization (follow-up). The aim was to keep concurrent medication stable during the optimization phase; however, psychiatrists were allowed to make changes for clinical indications (for an overview of psychotropic medications used over time see Supplementary Table 1). Healthy controls were scanned at two different time points (baseline and after 5 months follow-up). After the follow-up, the DBS group entered the double-blind randomized cross-over phase. This phase consisted of two blocks of one to six weeks during which DBS stimulation was on (active) or off (sham). Patients could be prematurely crossed over to the next phase (blinding was maintained), if this was requested by the patient or if the treating psychiatrist or research team deemed it clinically indicated and the HAM-D-17 score was at least 15. Concurrent medication and DBS settings were kept stable during the crossover phase. Patients again received MRI scans after active as well as sham stimulation. The severity of depressive symptoms was measured at each assessment (baseline, follow-up, active, sham) with the HAM-D-17 with higher scores indicating more severe depressive symptoms. Response to DBS was defined as a ≥50% reduction of HAM-D-17 score at follow-up compared to baseline. A description of the analyses of the HAM-D-17 scores can be found in Supplementary Methods 1.1.

### 2.4 fMRI Acquisition and Emotional Face Viewing Task

Structural and functional MRI data were collected with a 1.5T Siemens Magnetom Avanto syngo MR scanner with a transmit/receive (Tx/Rx CP) Head Coil. For safety reasons the DBS devices were switched off during scanning. For details on the acquisition parameters, see Supplementary Methods 1.2. To probe amygdala activation during fMRI data acquisition participants performed an implicit emotional face viewing task at each time point. Previous research has shown that looking at emotional faces consistently activates the amygdala (12). The task consisted of 36 blocks (6 for each condition) of 20 seconds (total time: 12 minutes). During each block participants were presented with four pictures (four seconds each) of a face with the same emotional expression (neutral, fearful, happy, angry or sad) or a scrambled face with an arrow pointing to the left or the right (control condition). Each block was concluded by a four second fixation cross. To ensure task attendance participants were asked to indicate the sex of the face (male or female) or to which direction the arrow was pointing (left or right) with a button press. Blocks had a pseudorandomized order and the same version was performed at each time point. The task was presented with E-prime (version 2.0; Psychology Software Tools, Inc.; Pittsburgh, PA). Reaction time and accuracy were recorded for each trial. A description of the analysis of the reaction times and accuracy can be found in Supplementary Methods 1.1.

### 2.4 fMRI Data Analysis

#### 2.4.1 Preprocessing

Before preprocessing a quality assessment was carried out by manually reviewing MRIQC (13) Image Quality Metrics and visual reports. (f)MRI data were preprocessed with MATLAB R2018b (The Math Works, Inc., 2018) using the SPM12 toolbox. Functional images were realigned to the mean functional image and subsequently slice-time corrected. Next, the T1-weighted image was coregistered to the mean functional image. The T1-weighted image was then segmented and forward deformation fields were created. Using these forward deformation fields, the functional images and structural image were normalized into standardized MNI space. To reduce noise, the functional images were smoothed using an 8-mm full width at half maximum Gaussian kernel.

#### 2.4.2 Amygdala and Whole-Brain Activity and Amygdala Functional Connectivity

Neural responsivity to faces was estimated with a first-level analysis for each participant at each time-point (baseline, follow-up, active and sham stimulation) using a general linear model with the emotional faces (neutral, fearful, happy, angry, sad), the control condition and the six realignment parameters modeled as separate regressors. Contrast images were created for each of the five emotional faces (face minus control) and were taken to the second-level group analyses. To assess amygdala responsivity, one region-of-interest (ROI) mask for the left and right amygdala was created based on the Talaraich Daemon atlas using the WFU PickAtlas tool embedded in SPM12. To investigate the effect of DBS on task specific functional connectivity with the rest of the brain we calculated psychophysiological interaction (PPI) functional connectivity, using the same amygdala ROI mask as seed region (for details, see Supplementary Methods 1.3).

#### 2.4.3 fMRI Statistical Analysis

All group-level fMRI analyses were performed using the Sandwich Estimator toolbox for SPM12 (SwE v2.2.0; http://www.nisox.org/Software/SwE) (14) (degrees of freedom type II, small sample adjustment for Wild Bootstrap resampling type C2, 999 bootstraps, unrestricted U-SwE). To assess DBS induced changes in amygdala responsivity and PPI functional connectivity from baseline compared to follow-up we performed separate mixed ANOVAs with group (DBS vs. healthy controls) as the between-subjects factor and session (baseline vs. follow-up) as the within-subjects factor. Emotional valence (neutral vs. fearful vs. happy vs. angry vs. sad) was additionally added as a within-subjects factor for the amygdala responsivity model. Since the time from baseline to follow-up varied greatly within patients (mean = 443 days, SD = 152.22) and between patients and healthy controls (mean = 149.25 days, SD = 14.18), time between sessions (in days) was added as a covariate to the models. To assess amygdala responsivity after active compared to sham stimulation, we carried out a repeated measures ANOVA with session (active vs. sham) and emotional valence (neutral vs. fearful vs. happy vs. angry vs. sad) as within-subjects factors. Due to premature cross-overs the time between sessions varied between patients (mean = 19 days, SD = 13.71). Therefore, the time between the two sessions (in days) as well as the randomization order were added as covariates to the model. To compare amygdala PPI functional connectivity after a period of active stimulation compared to sham stimulation, we carried out a repeated measures ANOVA with session (active vs. sham) as the within-subjects factor. The time between sessions (in days) and the randomization order were added as covariates to the model.

For the ROI-analyses, we used the SwE non-parametric Wild Bootstrap procedure with voxel-wise inference and statistical tests across the left and right amygdala were corrected for multiple comparisons using a false discovery rate (FDR) small volume correction. Additionally, to examine more widespread brain activation and amygdala functional connectivity related to processing of emotional faces we performed the same SwE non-parametric Wild Bootstrap procedure but with cluster-wise inference with a cluster-forming threshold of 0.01. Whole-brain statistical tests were corrected for multiple comparison using the family-wise error rate (FWER). Corrected p-values ≤ 0.05 were considered significant.

## 3. Results

In total, twenty-five patients and twenty-two healthy controls were included in the clinical trial. Complete baseline and follow-up fMRI data for the implicit emotional face viewing task were available for twelve patients and seventeen healthy controls (for the reasons for missing data, see Supplementary Table 2). Data from one additional patient were excluded because of insufficient brain coverage, and data from one healthy control were excluded after data quality control (see Supplementary Results 2.1). Hence, the final sample for the baseline/follow-up analyses consisted of eleven patients (6 responders/5 non-responders) and sixteen healthy controls. Response status for one patient was manually adjusted from non-responder to responder as the patient had been in remission over the course of the optimization and the follow-up HAM-D-17 score was inflated by depression unrelated physical complaints. Complete fMRI data during the cross-over phase were available for thirteen patients (7 responders/ 6 non-responders; for the reasons for missing data, see Supplementary Table 2).

### 3.1 Clinical Results

Demographic and clinical characteristics are shown in Table 1. Patients and healthy controls did not differ significantly in sex, age and estimated IQ. As expected, patients scored significantly higher on baseline measures of depressive symptom severity scores (HAM-D-17, MADRS and IDS-SR). There was a significant reduction in HAM-D-17 score from baseline to follow-up (baseline: mean = 23.82, 95%CI = 19.95-27.69; follow-up: mean = 15.55, 95%CI = 9.31-21.78; t=-2.680, p<0.05), and patients had a significantly lower HAM-D-score after active stimulation compared to sham stimulation (active: mean =14.92, 95%CI = 10.62-19.23; sham: mean = 22.54, 95%CI = 19.46-25.54; t=-3.200, p<0.05).

**Table 1.** Descriptive characteristics of DBS patients and healthy controls.

| Characteristics | Baseline – follow-up |  | p-value<sup>†</sup> | Active-sham |
| --- | --- | --- | --- | --- |
|  | DBS | HC |  | DBS |
| Sample size | 11 | 16 |  | 13 |
| **Sex, No. (%)** |  |  |  |  |
| Female | 10 (90.91) | 10 (62.5) | 0.227 | 8 (61.54) |
| Male | 1 (9.09) | 6 (37.5) |  | 5 (38.46) |
| Age at inclusion, mean (SD), y | 50.55 (7.88) | 53.56 (8.45) | 0.182 | 52.15 (7.77) |
| Estimated IQ, mean (SD) | 94.45<br>(16.21) | 103.13 (13.67) | 0.199 | 96.77 (15.57) |
| Baseline HAM-D-17 score, mean (SD) | 23.82 (5.76) | 0.88 (1.36) | <0.001* | 22 (5.94) |
| Baseline MADRS score, mean (SD) | 34.91 (6.67) | 0.88 (1.31) | <0.001* | 33.38 (6.33) |
| Baseline IDS-SR score, mean (SD) | 49.64 (9.81) | 3.38 (2.70) | <0.001* | 47.23 (11.83) |
| **Age at TRD onset, mean (SD), y** |  |  |  |  |
| Self-report | 26.36<br>(17.22) |  |  | 30.46 (14.40) |
| Diagnosis | 36 (11.04) |  |  | 37.23 (11.45) |
| No. of past medications, mean (SD) | 11.18 (3.60) |  |  | 10.46 (3.13) |
| No. of past ECT series, mean (SD) | 1.91 (1.04) |  |  | 1.85 (1.07) |
| No. of past ECT sessions, mean (SD) | 53.82<br>(54.61) |  |  | 46.92 (51.79) |
| Years since TRD onset, diagnosis, mean (SD) | 14.55 (7.93) |  |  | 14.92 (10.15) |
| **TRD episodes, No. (%)** |  |  |  |  |
| 1 | 6 (55.5) |  |  | 7 (53.8) |
| 2 | 2 (18.2) |  |  | 3 (23.1) |
| >2 | 3 (27.3) |  |  | 3 (23.1) |

*Abbreviations:* DBS, Deep Brain Stimulation; HC, healthy controls; SD, standard deviation; IQ, intelligence quotient; HAM-D, Hamilton Rating Scale for Depression; MADRS, Montgomery-Åsberg Depression Rating Scale; IDS-SR, Inventory of Depressive Symptomatology–Self-report; TRD, treatment-resistant depression; ECT, electroconvulsive therapy.
† p-values were obtained with appropriate parametric and non-parametric tests.
\* Significant differences between patients and healthy controls (p<0.001).

### 3.2 Behavioral Results

Two healthy controls and one patient did not press a button for more than half of the trials of the implicit emotional face viewing task during one of their sessions and were therefore excluded from the behavioral analyses. The final sample for the reaction time analyses included 14 healthy controls and 11 DBS patients (baseline vs. follow-up), and 12 DBS patients (active vs. sham). For the accuracy analysis one additional patient was excluded from the baseline/follow-up comparison due to an extreme outlier. There was a significant main effect of group for reaction time, with healthy controls being faster than DBS patients (F(1,22)=13.864, p=0.001). Additionally, there was a significant interaction between group and session (F(1,22)=4.301, p=0.05). Follow-up testing showed that patients became significantly faster at follow-up compared to baseline, whereas healthy controls performed virtually with the same speed during each session. There were no significant differences in accuracy. There were also no significant differences in reaction time or accuracy between active and sham stimulation (p>0.05).

### 3.3 Amygdala Responsivity

#### 3.3.1 Baseline vs. Follow-up

Whole-brain comparisons between all patients and healthy controls did not yield any significant clusters (p_FWE-corrected_>0.05). ROI-analysis of amygdala responsivity revealed a significant group x session interaction in the right amygdala (p_FDR-corrected_=0.017), but no significant group x session x valence interaction. Thus, the change in the right amygdala responsivity over time was different between healthy controls and DBS patients, but did not depend on emotional valence. Post-hoc comparisons showed significantly reduced right amygdala responsivity in patients compared to healthy controls at baseline (p_FDR-corrected_=0.018), but no significant difference in right amygdala activation at follow-up (p_FDR-corrected_>0.05) (figure 1). Additionally, right amygdala responsivity in healthy controls tended to decrease at follow-up compared to baseline (right: p_FDR-corrected_=0.077), indicating that DBS prevented amygdala habituation over time.

**Figure 1.**
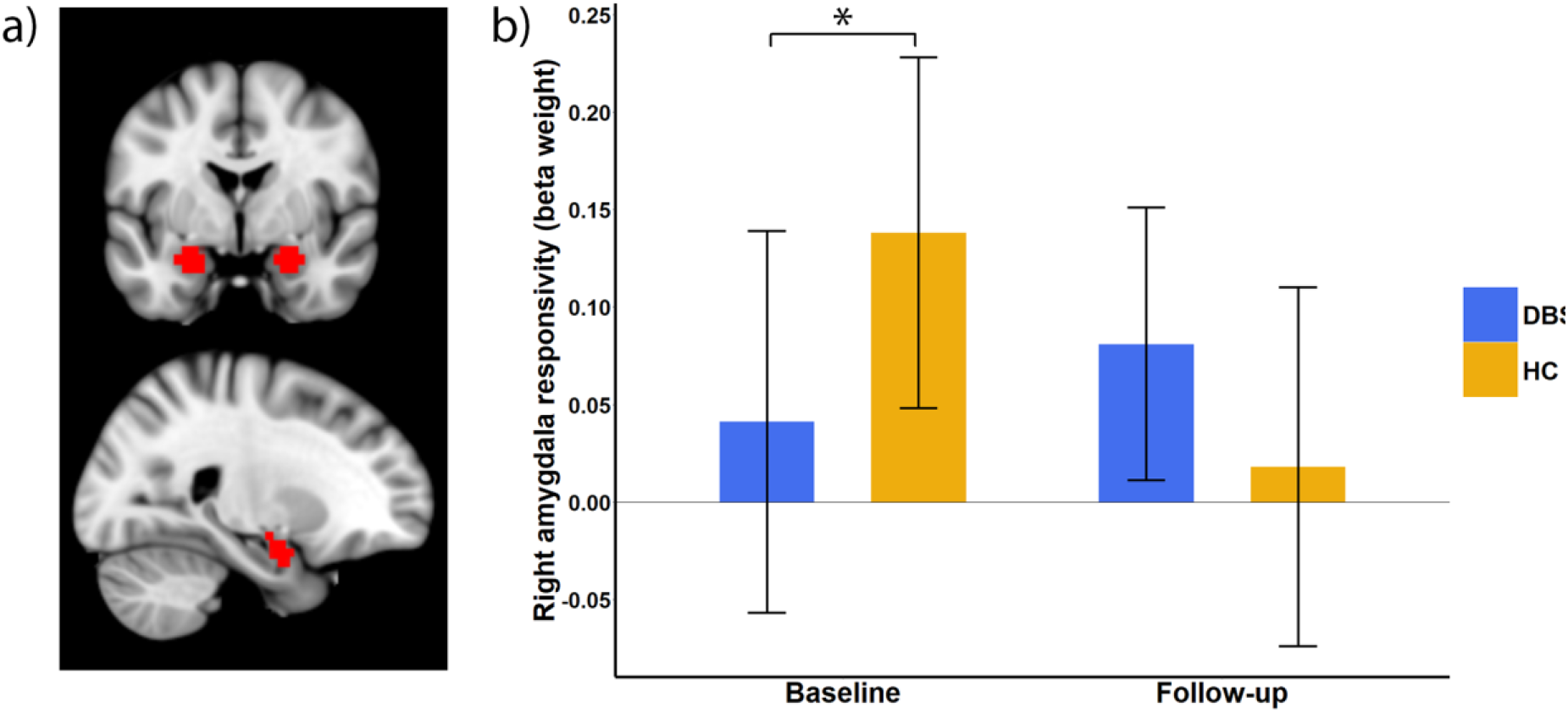
vALIC DBS normalizes amygdala hyporesponsivity. (a) Amygdala region-of-interest (ROI; red). (b) vALIC DBS-induced changes in right amygdala responsivity (mean with 95% CI); group × session interaction (p_FDR-corrected_=0.017). DBS patients show significantly reduced right amygdala activity at baseline compared to healthy controls (p_FDR-corrected_=0.018) but not at follow-up (p_FDR-corrected_>0.05).

For exploratory purposes we repeated the analyses but divided the patient group into responders and non-responders and compared them to the healthy controls. The ROI-analysis showed a trend towards a group x session interaction in the right amygdala (p_FDR-corrected_=0.069). Post-hoc comparisons showed significantly reduced right amygdala responsivity in responders compared to healthy controls at baseline (p_FDR-corrected_=0.044), but no significant difference in right amygdala responsivity in non-responders compared to healthy controls at baseline (p_FDR-corrected_>0.05). Compared to healthy controls neither the responders nor the non-responders showed a significant difference in right amygdala responsivity at follow-up (p_FDR-corrected_>0.05).

Inspection of fMRI data showed that significant results in the right amygdala did not overlap with BOLD-signal drop-out related to DBS electrodes (see Supplementary Figure 1).

#### 3.3.2 Active vs. Sham DBS

Whole-brain analysis and amygdala ROI-analysis of the cross-over phase (active vs. sham) showed no significant session x valence interaction effects and no significant main effects of session. Exploratory analyses with the patient group divided into responders and non-responders did not show significant session x group x valence or session x group interaction effects nor significant session or group main effects.

### 3.4 Functional Connectivity

#### 3.4.1 Baseline vs. Follow-up

Since the baseline/follow-up analysis only showed significant differences in the right amygdala, we used the right amygdala as the seed region for the PPI connectivity analyses (faces vs. control). Comparing all patients with healthy controls showed no significant group x session interaction nor session or group main effects. Exploratory analyses with the patient group divided into responders and non-responders and comparing them to healthy controls did not yield significant group x session interaction or session/group main effects either.

#### 3.4.2 Active vs. Sham DBS

PPI connectivity analysis (faces vs. control) showed increased connectivity during active compared to sham stimulation between the right amygdala and a large cluster in the left precentral gyrus (BA6) which also included the left postcentral gyrus (BA8/5), superior parietal cortex (BA7), inferior parietal cortex (BA40), right and left midcingulate region (BA24), right and left posterior cingulate cortex (BA23/30), and the left anterior cingulate cortex (ACC; BA24) (p_FWE-corrected_=0.027; Table 2; Figure 2). Exploratory analyses with the patient group divided into responders and non-responders did not show significant group x session interaction or session/group main effects. Inspection of fMRI data showed that significant results were not located in brain regions with BOLD-signal drop-out related to DBS electrodes (see Supplementary Figure 1).

**Table 2.** Peak coordinates of task specific right amygdala functional connectivity: active vs. sham DBS.

| PPI seed ROI | Connected region | Side | Cluster voxels | Peak-level MNI coordinates (x,y,z) |
| --- | --- | --- | --- | --- |
| *active>sham* |  |  |  |  |
| Right amygdala | Precentral gyrus (BA6) | L | 2059 | -22, -20, 60 |
|  | Midcingulate (BA24) | R |  | 4, -10, 44 |
|  | Posterior cingulate cortex (BA23/30) | L |  | -8, -40, 20 |

*Abbreviations:* DBS, deep brain stimulation; PPI, psychophysiological interaction; ROI, region-of-interest; BA, Brodmann area; L, left; R, right.

**Figure 2.**
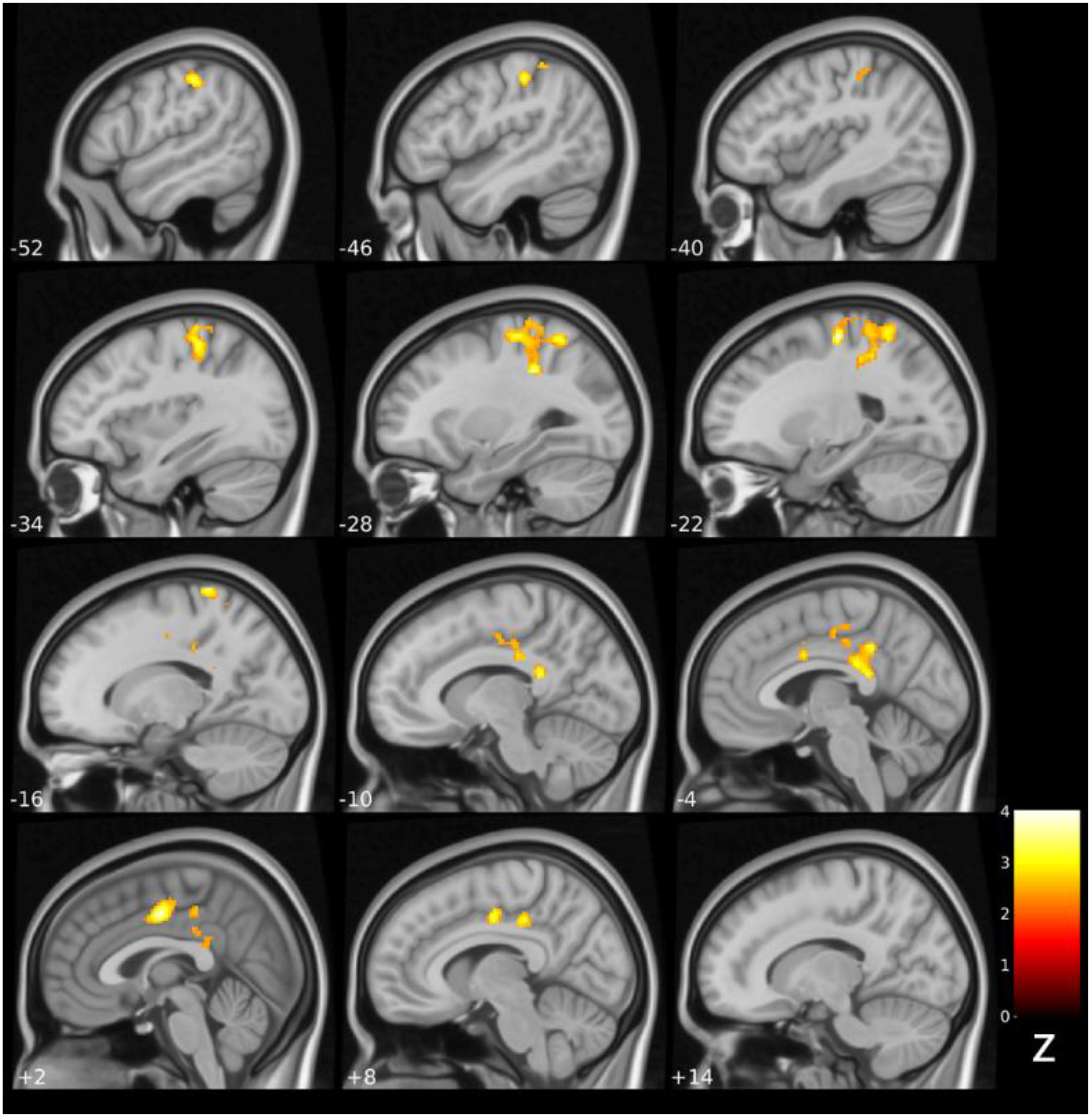
Statistical maps showing PPI functional connectivity after active compared to sham DBS (seed: right amygdala). Active vALIC DBS significantly increases functional connectivity between the right amygdala and a large cluster extending into the left precentral gyrus (BA6), postcentral gyrus (BA8/5), superior parietal cortex (BA7), inferior parietal cortex (BA40), right and left midcingulate region (BA24), right and left posterior cingulate cortex (BA23/30), and the left anterior cingulate cortex (ACC; BA24) compared to sham DBS (p_FWE-corrected_=0.027).

To determine if the difference in PPI connectivity between active and sham stimulation was task-dependent we performed an additional seed-voxel analysis of right amygdala connectivity irrespective of task condition. This analysis revealed no significant differences in connectivity between the right amygdala and the rest of the brain after active compared to sham stimulation (p_FWE-corrected_>0.05), indicating that the above-mentioned connectivity differences between active and sham stimulation were task-dependent.

### 3.5 Sensitivity Analyses

The neuroimaging analyses were repeated after exclusion of participants without sufficient behavioral data, which resulted in comparable results (see Supplementary Results 2.2).

## 4. Discussion

The aim of the present study was to determine how vALIC DBS affects amygdala responsivity and connectivity during an implicit emotional face viewing task. At baseline, TRD patients had moderate to severe depressive symptoms and showed decreased right amygdala responsivity compared to healthy controls. vALIC DBS resulted in a significant decrease in depressive symptoms and normalization of right amygdala responsivity. Active vALIC stimulation in comparison to sham stimulation was associated with increased task-dependent functional connectivity between the right amygdala and sensorimotor and cingulate cortices. Together, this indicates that the initial effect of vALIC DBS is associated with a change in right amygdala responsivity but not connectivity, whereas short-term effects of active vs. sham stimulation (after DBS parameter optimization) are related to right amygdala connectivity but not responsivity.

In line with studies showing DBS induced normalization of pathological brain activity (15-17), we found that vALIC DBS in TRD patients restored blunted right amygdala responsivity to healthy control levels. Although right amygdala responsivity in patients did not significantly increase over time, it tended to decrease in healthy controls, suggesting that repeated performance of the implicit emotional face viewing task led to amygdala habituation. Therefore, these results suggest that vALIC DBS resulted in a relative increase in right amygdala responsivity, which is in line with a previous finding of increased amygdala glucose metabolism after one week of NAc DBS (6). Additionally, we found that compared to healthy controls there was a significant decrease in reaction time during the implicit emotional face viewing task in patients after vALIC DBS compared to baseline. This suggests that the induced relative increase in right amygdala responsivity is accompanied by an increase in vigilance, which is in line with research showing that the amygdala plays an important role in vigilance modulation (18).

Our finding of decreased right amygdala responsivity at baseline is in contrast with previous findings of increased amygdala responsivity in MDD (19, 20). However, as a subtype of MDD TRD may be associated with different brain aberrancies (21). For example, one study found decreased amygdala responsivity in TRD patients (22) which may indicate emotional blunting. Although emotional blunting is frequently reported as a side effect of antidepressants (23), it is also thought to be a residual symptom of depression after antidepressant treatment (24). Together, this suggests that vALIC DBS may relieve depressive symptoms by mitigating emotional blunting through normalization of amygdala responsivity. On the other hand, amygdala responsivity in TRD is highly variable. Other studies found no difference or even increased amygdala responsivity compared to healthy controls (25, 26). Our exploratory analysis shows that only DBS responders have blunted right amygdala responsivity at baseline, which is subsequently increased by DBS (relative to healthy controls). Although this effect did not reach statistical significance, it is in line with a previous study that reported a significant increase in glucose metabolism in the right amygdala in responders compared to non-responders after 12 months of NAc DBS (4). Therefore, we speculate that variability in amygdala responsivity might be related to the effectiveness of DBS. However, further research is necessary to determine if baseline amygdala responsivity is a potential predictor of response for (vALIC) DBS.

Interestingly, the normalization of right amygdala responsivity following vALIC DBS did not depend on the valence of the emotional faces, suggesting that DBS influences amygdala responsivity to salient stimuli in general. This is in line with the hypothesis that the amygdala does not only process emotional expressions but plays a general role in the detection of and regulating the attention to salient or motivationally relevant stimuli (27). DBS may increase attention towards (emotionally) salient stimuli and have beneficial effects in settings that require additional attentional resources such as novel and uncertain situations. However, our study might have been underpowered to detect subtle differences between emotional valences.

In addition to the localized effects of DBS on the amygdala, vALIC DBS also increased task-dependent right amygdala connectivity with the precentral gyrus, postcentral gyrus, superior and inferior parietal cortex, midcingulate region, ACC, and posterior cingulate cortex. Although we did not find baseline differences in amygdala connectivity, a previous study reported decreased amygdala connectivity with the dorsal ACC in MDD in a similar location to our ACC cluster (28). This indicates that amygdala-ACC connectivity changes into a pathological state when DBS is switched off. The dorsal ACC is known as the cognitive division and is involved in modulation of attention or executive functions by influencing response selection, motivation and novelty (29). In addition, this cluster also included the posterior cingulate cortex, which has been linked to emotional salience processing independent of valence (30). Together, this further suggests that the effect of DBS may be associated with regulating attention towards (emotionally) salient stimuli. A diffusion tensor imaging study demonstrated structural connections between the amygdala and the pre- and postcentral cortex and midcingulate cortex (31). This direct amygdala-motor pathway may provide a mechanism by which amygdala processes can influence motor behaviors in (emotionally) salient contexts. Thus, the effects of DBS on amygdala-motor cortex connectivity may have an impact on motor behavior.

A particular strength of our study was the inclusion of a healthy control group, which allowed us to detect DBS induced changes adjusted for effects of repeated testing. The main limitation of the study is the small sample size, even though this study is the largest to date to investigate the neural effects of DBS in MDD. This study was part of a randomized controlled clinical trial for which the initial sample size was determined to have sufficient power to detect the therapeutic efficacy of vALIC DBS (3), and for which the effects of DBS on the brain were secondary outcomes. Unfortunately, complete fMRI data was only available for about half of the included patients. The results nevertheless show that the sample size was sufficient to obtain results that remained significant after correction for multiple voxel-wise comparisons.

In conclusion, the strong effect of vALIC DBS on depressive symptoms was associated with normalization of blunted baseline amygdala responsivity, as well as an increase in vigilance as reflected by faster reaction times. In addition, active stimulation increased task-dependent amygdala connectivity with cingulate and sensorimotor regions compared to sham stimulation. Together, these results suggest that vALIC DBS improves depressive symptoms in part by reversing amygdala pathology in MDD.

## Data Availability

All data produced in the present study are available upon reasonable request to the authors

## Acknowledgments

The authors would like to thank Frank van Thienen for his comments and suggestions on the grammar and punctuation of the manuscript.

Funding: This investigator-initiated study was funded by Medtronic Inc (25 DBS systems, in kind) and a grant from ZonMw (nr. 171201008).

