## Supplementary information for "Deep Brain Stimulation Normalizes Amygdala Responsivity in Treatment-Resistant Depression"

*Content:*

1. *Supplementary Methods*
    - *1.1 Clinical and behavioral statistical analyses*
    - *1.2 MRI Acquisition Parameters*
    - *1.3 PPI Functional Connectivity*
2. *Supplementary Results*
    - *2.1 Quality Control Based Exclusions*
    - *2.2 Sensitivity Analyses*

*Supplementary Table 1*

*Supplementary Table 2*

*Supplementary Figure 1*

### *1.* Supplementary Methods

#### *1.1 Clinical and behavioral statistical analyses*

Clinical and behavioral data were analyzed with R version 4.0.3 (R Core Team, 2020). HAM-D-17 scores at baseline and follow-up were compared with a linear mixed-effects model analysis with HAM-D-17 score as the dependent variable and session (baseline vs. follow-up) and time between sessions as independent variables. In addition, for the active/sham stimulation comparison we performed a linear mixed-effects model analysis with HAM-D-17 as the dependent variable and session (active vs. sham), time between sessions, and randomization order as independent variables.

Reaction time and accuracy during the implicit emotional face viewing task at baseline and follow-up were analyzed using two separate mixed ANOVAs with group (DBS vs. healthy controls) as the between-subjects factor, session (baseline vs. follow-up) as the within-subjects factor, and time to follow-up as a covariate. Additionally, for the active/sham stimulation comparison we performed two linear mixed-effects model analyses with reaction time and accuracy as the dependent variables respectively, and session (active vs. sham), time between sessions, and randomization order as independent variables.

#### *1.2 MRI Acquisition Parameters*

Echo-planar images with transversal orientation were acquired during the implicit emotional face viewing task (repetition time=2000 ms, echo-time=30 ms, flip angle=90°, matrix=64x64, number of slices=25, slice thickness=4.0 mm, slice gap=10%, slice order=interleaved ascending (odd first), field of view=230x230, voxel-size= 3.6x3.6x4.0 mm). 360 volumes were acquired with a total duration of 12 minutes. For anatomical reference, a three dimensional single shot T1-weighted image with sagittal orientation was acquired (repetition time=1900 ms, echo-time=3.08 ms, flip angle=15°, matrix =512x512, number of slices=192, slice thickness=1.0 mm, slice gap=50%, field of view=256x256, voxel-size=0.5x0.5x1.0 mm).

#### 1.3 PPI Functional Connectivity

To calculate the PPI between the amygdala and the rest of the brain, the amygdala bold-response was extracted from the original first-level models (see main text) adjusted for the effects of interest contrast. Using the original first-level models and the extracted amygdala BOLD-response, the PPI was calculated using SPM12. For this calculation each of the emotional faces (neutral, fearful, happy, angry, sad) were assigned a contrast weight of 1, and the control condition was assigned a contrast weight of -1. Afterwards, a new set of first-level models were created with the PPI and the six realignment parameters modeled as separate regressors. PPI connectivity contrast images were generated from the respective first-level models and taken to the second-level group analyses.

### 2. Supplementary Results

#### 2.1 Quality Control Based Exclusions

fMRI data from one patient and two healthy controls were identified as being of potentially insufficient quality after reviewing the MRIQC (1) Image Quality Metrics and visual reports. Subsequently, we performed positive control analyses and for one healthy control we did not observe visual or motor cortex activity at first level contrasts (visual contrast: faces and control vs. fixation cross; motor contrast: button press vs. fixation cross). We therefore excluded this healthy control from the baseline and follow-up fMRI analyses.

#### 2.2 Sensitivity Analyses

Two healthy controls and one patient did not press a button for more than half of the trials of the implicit emotional face viewing task during one of their sessions. These participants may not have properly attended to the emotional faces during fMRI data acquisition. To ensure that this did not influence our results, we repeated all fMRI analyses while excluding these participants. The main results remained the same (i.e. significant results remained significant ($p_{corrected}<0.05$) and non-significant results remained non-significant ($p_{corrected}>0.05$)). Results were only different for one of the exploratory analyses where we divided the patient group into responders and non-responders and compared them to the healthy controls: there no longer was a trend towards an interaction between the effects of group (responder vs. non-responder vs. healthy controls) and session (baseline vs. follow-up) on amygdala responsivity.

*Supplementary Table 1*

| Supplementary Table 1. Number of patients using psychotropic medication over time | | | | | |
|---|---|---|---|---|---|
| | | Baseline – follow-up (n=11) | | Active-sham (n=13) | |
| | | Baseline | Follow-up | Baseline | Follow-up |
| **Antidepressant** | Combination | 1 | 2 | 1 | 3 |
| | Single | 6 | 2 | 6 | 3 |
| | None | 4 | 7 | 6 | 7 |
| **Benzodiazepine** | Combination | 1 | 0 | 0 | 0 |
| | Single | 4 | 5 | 5 | 5 |
| | None | 6 | 6 | 8 | 8 |
| **Antipsychotic** | Single | 6 | 6 | 6 | 5 |
| | None | 5 | 5 | 7 | 8 |
| **Lithium** | Single | 1 | 0 | 1 | 1 |
| | None | 10 | 11 | 12 | 12 |
| **Anxiolytic** | Single | 0 | 1 | 0 | 1 |
| | None | 11 | 10 | 13 | 12 |
| **Anti-epileptic** | Single | 1 | 0 | 1 | 1 |
| | None | 10 | 11 | 12 | 12 |
| **Antihistaminic** | Single | 1 | 1 | 1 | 1 |
| | None | 10 | 10 | 12 | 12 |
| **Opioid** | Single | 0 | 0 | 1 | 1 |
| | None | 11 | 11 | 12 | 12 |
| **Sympathomimetic** | Single | 1 | 1 | 2 | 2 |
| | None | 10 | 10 | 11 | 11 |

*Supplementary Table 2*

**Supplementary Table 2. Reasons for missing fMRI data.**

| **Patient** | **Complete baseline/ follow-up fMRI data** | **Complete cross-over phase fMRI data** | **Reason for missing data** |
|---|---|---|---|
| 1 | No | No | MRI coil was unavailable at baseline. Because there was no baseline data it was decided to not collect fMRI data at follow-up and during the cross-over phase. However, from this patient onwards fMRI data was collected at the following assessments despite missing baseline data. |
| 2 | No | Yes | MRI coil was unavailable at baseline. |
| 3 | No | Yes | MRI coil was unavailable at baseline. |
| 4 | Yes | Yes | |
| 5 | Yes | No | Patient was deemed unfit to participate in the cross-over phase due to unstable clinical status. |
| 6 | No | No | Drop-out due to non-response. |
| 7 | No | No | Patient was treated with MRI-incompatible vagus nerve stimulation. |
| 8 | No | No | Drop-out due to non-response. |
| 9 | Yes | Yes | |
| 10 | No | No | Drop-out due to non-response. |
| 11 | Yes | Yes | |
| 12 | No | No | Follow-up time deviated too much from protocol (2.5 years). Patient was deemed unfit to participate in the cross-over phase due to unstable clinical status. |
| 13 | Yes | Yes | |
| 14 | No | No | Patient withdrew from participation after the baseline assessment due to somatic complaints. |
| 15 | Yes | Yes | |
| 16 | No | No | Drop-out due to non-response. |
| 17 | Yes | Yes | |
| 18 | Yes | No | Unknown |
| 19 | No | Yes | Baseline fMRI data was not collected due to back complaints at the time. |
| 20 | Yes | No | fMRI data collection was terminated due to an anxiety attack during one of the cross-over assessments. |
| 21 | Yes | Yes | |
| 22 | No | Yes | At follow-up the buttons necessary for the fMRI task did not work. |
| 23 | No | Yes | fMRI data collection was terminated due to an anxiety attack at the follow-up assessment |
| 24 | Yes | Yes | |
| 25 | Yes | No | Patient was deemed unfit to participate in the cross-over phase due to unstable clinical status. |

| **Healthy control** | **Complete baseline/ follow-up fMRI data** | | **Reason for missing data** |
|---|---|---|---|
| 1 | Yes | | |
| 2 | Yes | | |
| 3 | Yes | | |
| 4 | Yes | | |
| 5 | Yes | | |
| 6 | Yes | | |
| 7 | Yes | | |
| 8 | No | | Participant withdrew from participation after the baseline assessment |
| 9 | Yes | | |
| 10 | Yes | | |
| 11 | Yes | | |
| 12 | Yes | | |
| 13 | No | | Unknown |
| 14 | Yes | | |
| 15 | No | | Unknown |
| 16 | No | | MRI-incompatible elbow pin |
| 17 | No | | fMRI data collection was terminated due to an anxiety attack |
| 18 | Yes | | |
| 19 | Yes | | |
| 20 | Yes | | |
| 21 | Yes | | |
| 22 | Yes | | |

*Abbreviations:* (f)MRI, (functional) magnetic resonance imaging;

*Supplementary Figure 1*

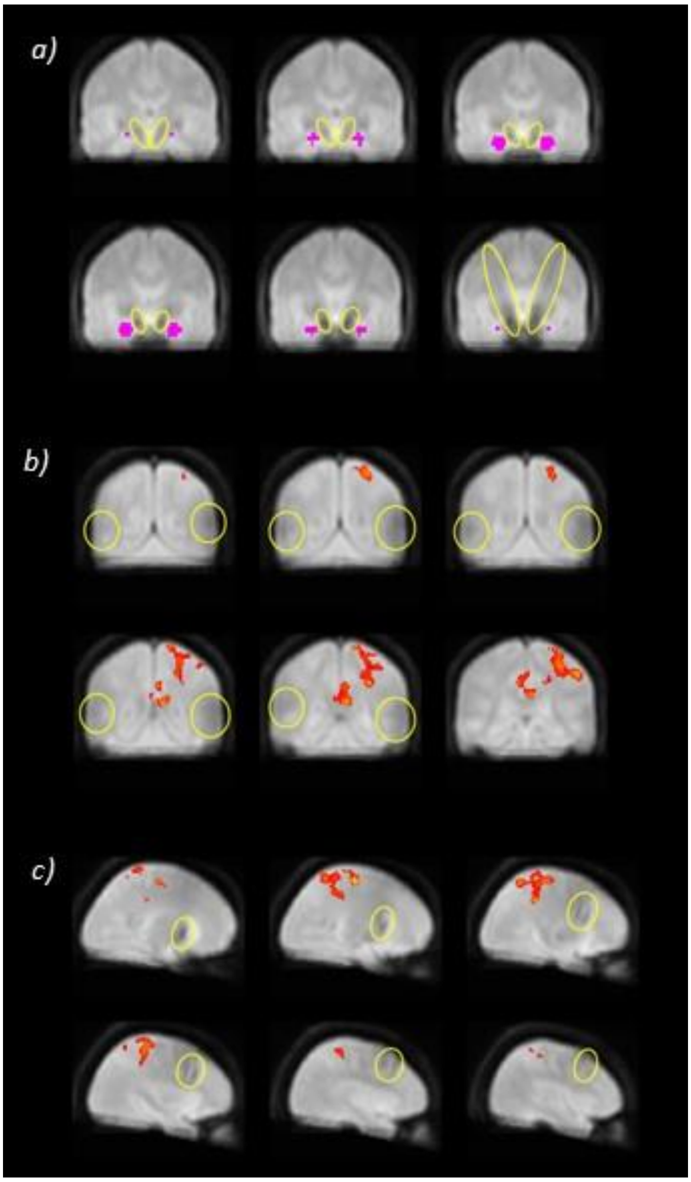


**Supplementary Figure 1. Mean images of all the normalized mean functional images displaying BOLD signal drop-out related to the DBS electrodes or extension wires.** Significant results were not located in brain regions with BOLD signal drop-out related to DBS electrodes or extension wires. (a) BOLD signal drop-out related to DBS electrodes (outlined in yellow) relative to the amygdala ROI (purple). (b) BOLD signal drop-out related to the DBS extension wire (outlined in yellow) relative to the significant amygdala connectivity cluster (red). (c) BOLD signal drop-out related to DBS electrodes (outlined in yellow) relative to the significant amygdala connectivity cluster (red).
